# Older racial and ethnic minority patients’ perceptions of Medicare Annual Wellness Visits: a qualitative study

**DOI:** 10.1101/2025.09.19.25336216

**Authors:** Tiffany Brown, Tahniat Nadeem, Carla Salazar, Jeffrey A. Linder, Gayle Kricke, David T. Liss

**Author notes:** Corresponding author: David T. Liss, PhD. Funding statement: Funding support was provided by the National Institute on Aging (#R21 AG081895). The funder had no role in the design and conduct of the study; management, analysis, and interpretation of the data; preparation, review, or approval of the manuscript; or decision to submit the manuscript for publication. Acknowledgments: We wish to thank patients who served as focus group participants, as well as staff at participating practices where focus groups were held. We additionally wish to thank our partners at AllianceChicago for supporting data collection efforts during study recruitment. Conflicts of interest: The authors report no conflicts of interest or competing interests.

## Abstract

**Purpose:** Although Medicare Annual Wellness Visits (AWVs) offer many potential benefits to older adults, racial and ethnic minorities have lower rates of AWV completion. Our objective was to understand older minority patients’ attitudes and preferences related to preventive care and AWVs.

**Methods:** Between June and October 2024, we conducted four focus groups at two urban primary care settings—one academic health system, and one federally qualified health center—among Medicare enrollees aged ≥66 with ≥1 primary care encounter at a participating organization in the prior year. We recruited patients with Black race or Hispanic ethnicity as documented in the electronic health record. Domains of interest were communication preferences, attitudes about preventive care and AWVs, and barriers to care. Focus groups were audio recorded, and transcripts were coded into key themes using a template analysis approach.

**Results:** There were 45 participants, who had a mean age of 71 years (standard deviation, 4); most were female and identified as Black. Participants reported varying forms of preferred communication, including the patient portal, phone calls, and mailed letters. Five themes emerged: 1) The value placed on participants’ health and preventive care; 2) The value placed on relationships with trusted primary care physicians; 3) Barriers to scheduling and attending doctors’ visits; 4) Confusion or uncertainty about terminology describing preventive visits, and; 5) Lack of trust due to historical discrimination.

**Conclusions:** Interventions to increase Medicare AWV uptake in racial and ethnic minority patients must address and overcome barriers such as those identified here.

## INTRODUCTION

Checkup visits—i.e., general health checks, preventive visits—benefit patients in multiple ways.^1^ These visits may be especially beneficial in older adults; randomized trials in patients aged ≥65 years consistently demonstrated that checkups improve preventive services uptake and patient-reported outcomes,^2–5^ and in some instances reduced total mortality.^5,6^ In the United States, the Annual Wellness Visit (AWV) is Medicare’s form of the checkup. AWVs are available to all Medicare and Medicare Advantage enrollees once a year, at zero copay.

Despite the potential benefits of AWVs, quantitative analyses have shown low AWV completion in racial and ethnic minorities,^7,8^ particularly among Black patients,^9–11^ as well as patients with low income^7,8^ or low education level.^9,10^ At our institution, recent research identified additional characteristics associated with AWV noncompletion, including lower use of advanced patient portal functionalities, prior positive screen for social needs (e.g. food insecurity), and AWV noncompletion during the prior year.^8^ In a prior qualitative study among older adults, most of whom were Black and lived in an assisted living facility, participants generally reported that they valued preventive care but many did not know about AWVs or had trouble understanding the term “wellness visit.” Many patients were more familiar with terms such as “checkup” or “physical.”^12^

To leverage the full potential of AWVs to improve population health in at-risk groups, new strategies must be developed to reach patients who are unaware of AWVs or face barriers to AWV completion, tailor AWV delivery processes to their needs, and increase their rates of AWV completion. To that end, our team conducted focus groups among older community-dwelling, racial and ethnic minority patients at two urban primary care practices, with the goal of identifying factors that could inform development of a subsequent intervention to increase AWV completion in at-risk groups. Our specific aim was to describe perceptions of preventive care and preventive visits, knowledge of AWVs, and barriers to AWV completion among older racial and ethnic minority patients at urban primary care practices.

## METHODS

### Study Design and Setting

This study was conducted at two organizations in Chicago, Illinois. Northwestern Medicine (NM) is a large academic health system that has more than 60 primary care clinics in northeastern Illinois. Near North Health is a federally qualified health center (FQHC) with eight clinics throughout Chicago. Between June-October 2024, we conducted four focus groups with older adults. Two groups were held at an NM practice in downtown Chicago among English-speaking patients. Two groups were conducted at Near North Health practices, one in English (at a practice on Chicago’s South Side) and one in Spanish (at a practice on Chicago’s West Side). Focus groups were two hours in duration. Participants received a $75 gift card honorarium at the end of each focus group, and participants who drove to NM focus groups received a voucher for free parking.

Northwestern University’s Institutional Review Board was the single reviewing IRB for the study and approved study protocols.

### Participants

Patients were eligible for focus group participation if they were an adult age ≥66 years, had Medicare insurance, self-reported Black race or Hispanic ethnicity, and had and at least one primary care encounter at a participating organization in the prior 12 months. Patients had to have a preferred language of English or Spanish for participation in each respective language.

During recruitment, we identified eligible patients through queries of electronic data warehouses containing organizational electronic health record data. Among those meeting eligibility criteria, we randomly selected batched samples of potential participants and sent them a mailed letter describing the study and how they could opt out of future study communications. Patients who did not opt out received up to three recruitment phone calls; those agreeing to participate received scheduling information about their focus group’s date and location.

Participants provided written informed consent at the beginning of each in-person focus group. Prior to group discussion, each participant completed a paper survey on self-reported demographic characteristics, chronic illnesses, self-rated health,^14^ and a single-item health literacy screener.^15^ Participants also completed survey items on barriers to primary care visit attendance, and how they contacted their primary care clinician outside the office.

### Data Collection

The focus group moderator guide prompted participants to discuss the following domains: (1) Communication preferences about recommended preventive services; (2) Attitudes and thoughts on preventive visits in general and Annual Wellness Visits specifically, including terminology and key components of these encounters; (3) Barriers and challenges to scheduling and attending wellness visits, and; (4) Recommendations on how to describe these visits to patients.

Focus groups were audio recorded and transcribed. Transcriptions were checked for accuracy against the recording. Prior to coding, the Spanish group’s transcription was translated into English and checked for accuracy by a native Spanish-speaking study team member (CS).

### Analysis

Focus group data were analyzed using a template analysis approach,^16,17^ as our team has done previously.^18^ During data collection, we developed an initial code list using domains from the moderator guide. After data collection was completed, each transcript had line numbers added and unique segments identified. Coders then jointly refined the initial coding scheme to capture insights gleaned from transcripts. Among study team members who coded the transcripts (TB, TN, and CS), two coders independently coded each transcript segment into a primary domain category using a content analysis approach.^19^ Coders met jointly to resolve any discrepancies, with disagreements reconciled by a third coder as needed.

## RESULTS

There were a total of 45 participants across the 4 focus groups (Table 1), with 25 participants at the academic health system (all English speakers) and 20 at the FQHC (15 English speakers, 5 Spanish speakers). Over two-thirds of participants were female, and most participants were Black. Higher proportions of FQHC participants reported current Medicaid coverage (53%), as well as chronic illness diagnoses such as diabetes (50%), chronic obstructive pulmonary disease (28%), and asthma (28%). FQHC patients also reported barriers to primary care visit attendance at higher rates, including finding a time when an appointment is available (41%, versus 28% of academic health system patients), visit copays (25% versus 8%) and transportation barriers (24% versus 4%).

**Table 1:** Focus Group Participant Characteristics.

| Characteristic | Academic Health System | FQHC<sup>c</sup> |
| --- | --- | --- |
| *N* (Spanish speakers) | 25 (0) | 20 (5) |
| Age, mean (SD) | 72 (4) | 70 (4) |
| Sex |  |  |
| Female | 17 (68%) | 15 (75%) |
| Male | 8 (32%) | 5 (25%) |
| Race/ethnicity, n (%)<sup>a, b</sup> |  |  |
| Hispanic or Latino | 3 (12%) | 6 (32%) |
| Black or African American | 21 (84%) | 13 (68%) |
| Mixed/multiple | 1 (4%) | 0 (0%) |
| Medicaid coverage, n (%)<sup>a</sup> |  |  |
| Current | 4 (16%) | 10 (53%) |
| Previous | 2 (8%) | 6 (32%) |
| Chronic illnesses, n (%)<sup>a, b</sup> |  |  |
| Diabetes | 6 (24%) | 9 (50%) |
| COPD, chronic bronchitis or emphysema | 2 (8%) | 5 (28%) |
| Asthma | 1 (4%) | 5 (28%) |
| High blood pressure or hypertension | 19 (76%) | 14 (78%) |
| Coronary artery disease | 4 (16%) | 1 (6%) |
| Heart failure | 3 (12%) | 1 (6%) |
| Self-rated health, n (%)<sup>a</sup> |  |  |
| Poor | 2 (8%) | 0 (0%) |
| Fair | 5 (20%) | 5 (26%) |
| Good | 9 (36%) | 13 (68%) |
| Very good | 9 (36%) | 0 (0%) |
| Excellent | 0 (0%) | 1 (5%) |
| Self-rated health, 1-5 mean (SD)<sup>a</sup> | 3.0 (1.0) | 2.8 (0.7) |
| Limited health literacy, n (%)<sup>a</sup> | 2 (8%) | 4 (21%) |
| Barriers to primary care visit attendance, n (%)<sup>a, b</sup> |  |  |
| Hard to find a time when appointment available | 7 (28%) | 7 (41%) |
| Copays for medicines, bloodwork, or other tests | 4 (16%) | 4 (24%) |
| Visit copays | 2 (8%) | 4 (24%) |
| Fear/worry about what will happen at visit | 1 (4%) | 5 (29%) |
| Hard to walk or get around | 2 (8%) | 3 (18%) |
| Hard to find or pay for transportation | 1 (4%) | 4 (24%) |
| Hard to fill out forms at the visit | 0 (0%) | 3 (18%) |
| How contacts primary care clinician outside office visits, n (%)<sup>a, b</sup> |  |  |
| Email/Patient portal/MyChart messages | 19 (76%) | 1 (5%) |
| Phone calls | 18 (72%) | 16 (80%) |
| Video calls | 2 (8%) | 0 (0%) |

<sup>a</sup> Self-reported data collected via paper survey administered at beginning of each focus group
<sup>b</sup> Participants asked to select all that apply; responses not mutually exclusive
<sup>c</sup> Data missing for some individual survey items among FQHC focus group participants
Abbreviations: COPD, chronic obstructive pulmonary disease; FQHC, federally qualified health center; SD, standard deviation

### Patient Communication with Primary Care Team

In the participant survey and group discussions, there was heterogeneity regarding how participants communicated with their primary care clinicians. In the survey, most academic health system participants reported using the MyChart patient portal (Epic Systems; Verona, WI); reported portal use was lower among FQHC participants (Table 1). Majorities of participants at both organizations reported speaking with their primary care clinician or care team via phone calls.

In focus group discussions, participants stated a variety of preferences on how they would like to be contacted about recommended preventive services. Although most NM participants were registered patient portal users, they expressed varying beliefs on the portal’s value. Some participants expressed positive views about how “everything is there” in MyChart, while others reported being overwhelmed by the large amount of information and medical terminology they were exposed to during portal use. Several participants expressed a preference for phone calls, while others cited the value of text messages or mailed letters, due to the availability or ease of consuming these communications. For example, one participant said of mailings, “[Group 3] And you know, for me, if you just sent it through the mail, I got a paper copy. Okay, I got it. I don’t have to hear that phone ringing… I don’t have to stop what I’m doing… I prefer mail, because mail is silent. I can open it and you can bring it with you.” Additionally, several participants said they preferred to receive multiple reminders via two or more communication channels.

### Key Themes on Medicare AWVs

Five key themes emerged from focus group discussions. We report on each theme below; Table 2 contains additional illustrative quotes from participants. While the emergent themes were similar across the two primary care settings, a few differences between the academic health system site and FQHC site are notable. FQHC participants were less comfortable and familiar with using the patient portal. They also spent more time discussing Medicare Advantage-related issues, such as overlap between AWVs and in-home visits offered by payers. In the Spanish language group, language barriers and translation concerns emerged.

**Table 2:**
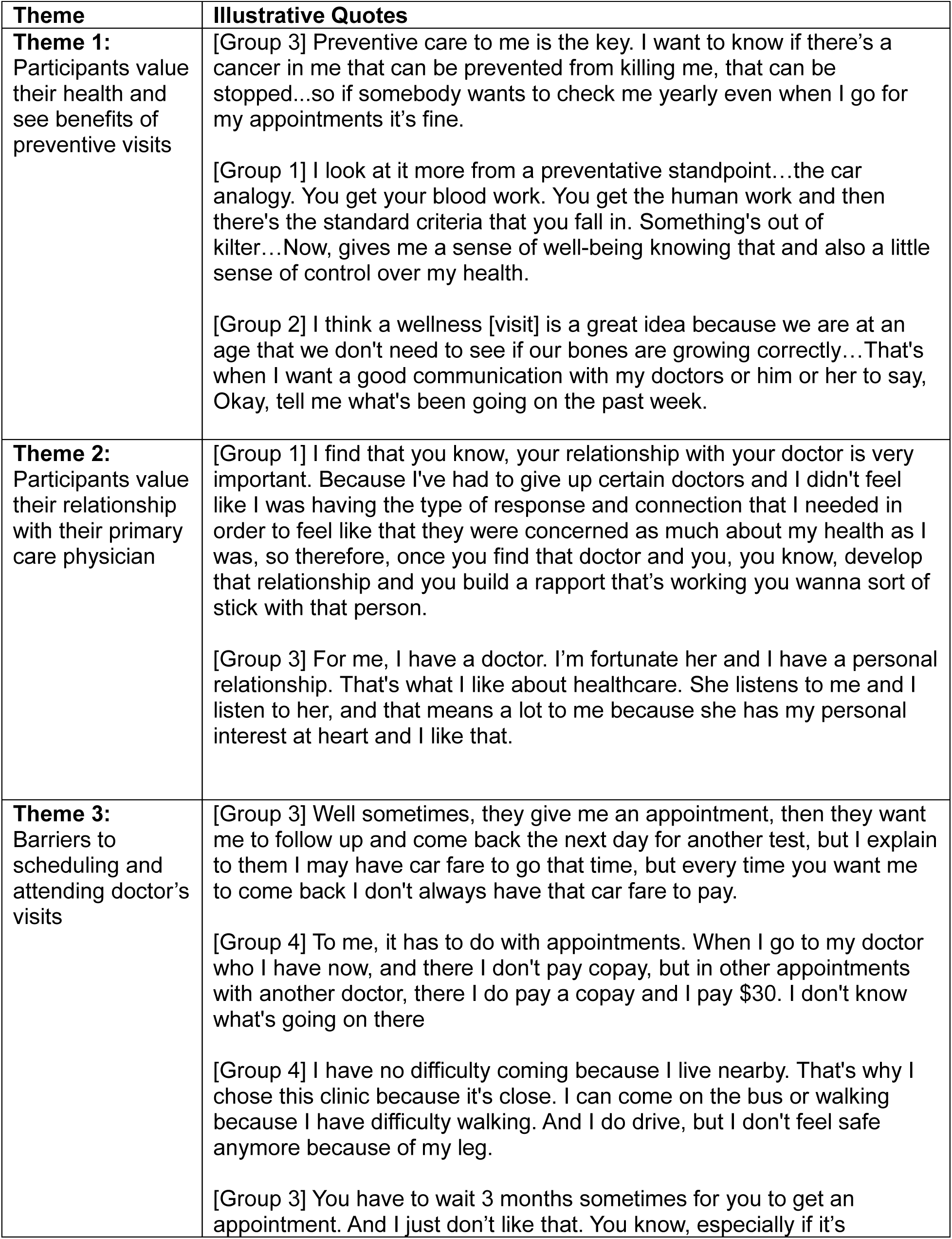
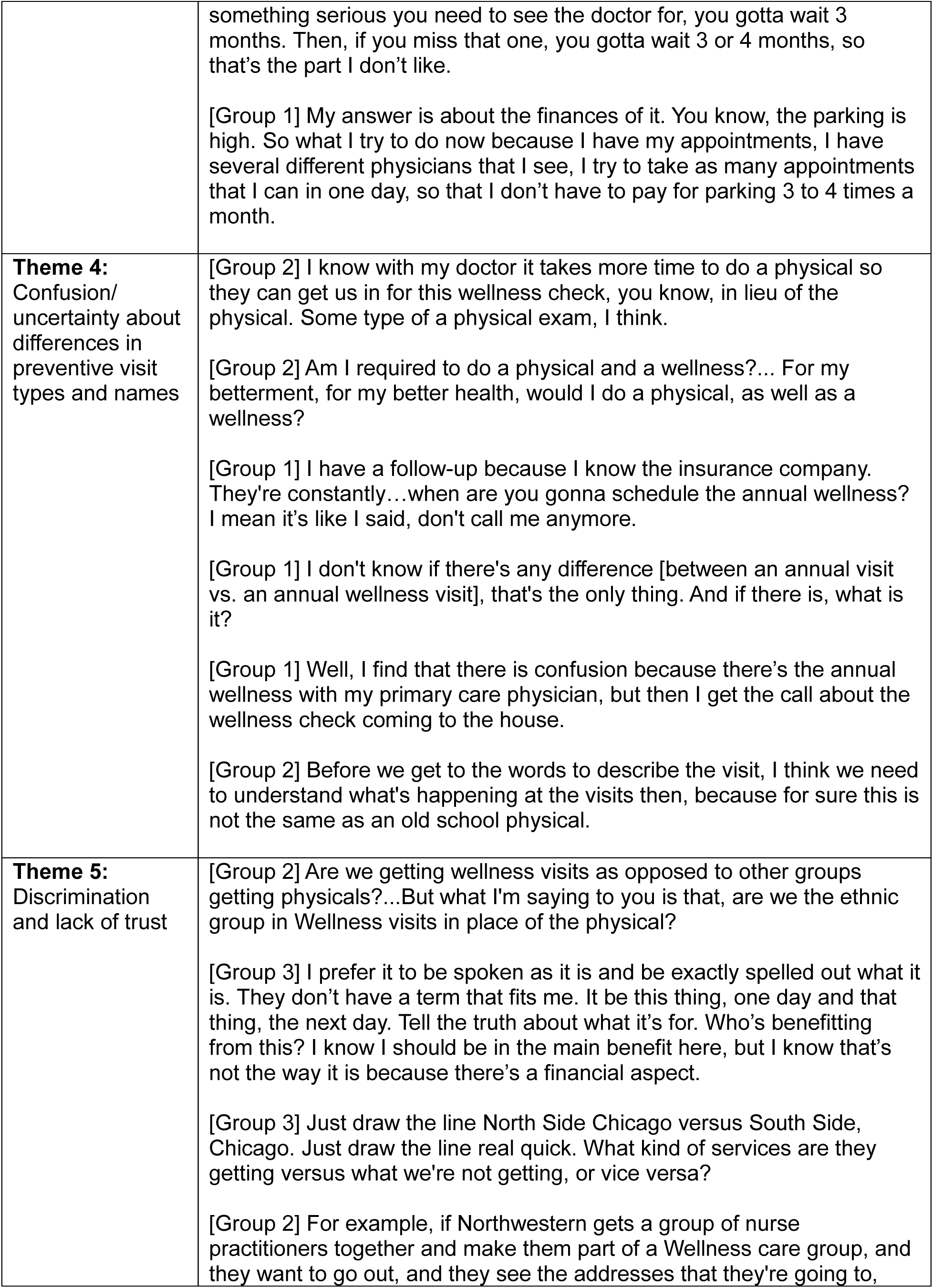
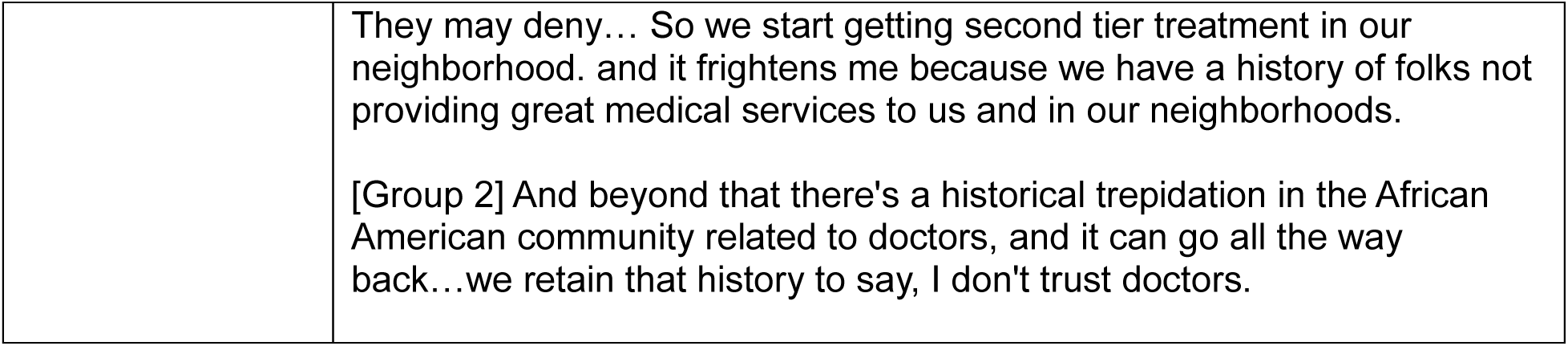
Key Themes and Illustrative Quotes.

| Theme | Illustrative Quotes |
| --- | --- |
| **Theme 1:**<br>Participants value their health and see benefits of preventive visits | [Group 3] Preventive care to me is the key. I want to know if there's a cancer in me that can be prevented from killing me, that can be stopped...so if somebody wants to check me yearly even when I go for my appointments it's fine.<br>[Group 1] I look at it more from a preventative standpoint...the car analogy. You get your blood work. You get the human work and then there's the standard criteria that you fall in. Something's out of kilter...Now, gives me a sense of well-being knowing that and also a little sense of control over my health.<br>[Group 2] I think a wellness [visit] is a great idea because we are at an age that we don't need to see if our bones are growing correctly...That's when I want a good communication with my doctors or him or her to say, Okay, tell me what's been going on the past week. |
| **Theme 2:**<br>Participants value their relationship with their primary care physician | [Group 1] I find that you know, your relationship with your doctor is very important. Because I've had to give up certain doctors and I didn't feel like I was having the type of response and connection that I needed in order to feel like that they were concerned as much about my health as I was, so therefore, once you find that doctor and you, you know, develop that relationship and you build a rapport that's working you wanna sort of stick with that person.<br>[Group 3] For me, I have a doctor. I'm fortunate her and I have a personal relationship. That's what I like about healthcare. She listens to me and I listen to her, and that means a lot to me because she has my personal interest at heart and I like that. |
| **Theme 3:**<br>Barriers to scheduling and attending doctor's visits | [Group 3] Well sometimes, they give me an appointment, then they want me to follow up and come back the next day for another test, but I explain to them I may have car fare to go that time, but every time you want me to come back I don't always have that car fare to pay.<br>[Group 4] To me, it has to do with appointments. When I go to my doctor who I have now, and there I don't pay copay, but in other appointments with another doctor, there I do pay a copay and I pay \$30. I don't know what's going on there<br>[Group 4] I have no difficulty coming because I live nearby. That's why I chose this clinic because it's close. I can come on the bus or walking because I have difficulty walking. And I do drive, but I don't feel safe anymore because of my leg.<br>[Group 3] You have to wait 3 months sometimes for you to get an appointment. And I just don't like that. You know, especially if it's |
|  | something serious you need to see the doctor for, you gotta wait 3 months. Then, if you miss that one, you gotta wait 3 or 4 months, so that's the part I don't like.<br>[Group 1] My answer is about the finances of it. You know, the parking is high. So what I try to do now because I have my appointments, I have several different physicians that I see, I try to take as many appointments that I can in one day, so that I don't have to pay for parking 3 to 4 times a month. |
| **Theme 4:**<br>Confusion/<br>uncertainty about<br>differences in<br>preventive visit<br>types and names | [Group 2] I know with my doctor it takes more time to do a physical so they can get us in for this wellness check, you know, in lieu of the physical. Some type of a physical exam, I think.<br>[Group 2] Am I required to do a physical and a wellness?... For my betterment, for my better health, would I do a physical, as well as a wellness?<br>[Group 1] I have a follow-up because I know the insurance company. They're constantly...when are you gonna schedule the annual wellness? I mean it's like I said, don't call me anymore.<br>[Group 1] I don't know if there's any difference [between an annual visit vs. an annual wellness visit], that's the only thing. And if there is, what is it?<br>[Group 1] Well, I find that there is confusion because there's the annual wellness with my primary care physician, but then I get the call about the wellness check coming to the house.<br>[Group 2] Before we get to the words to describe the visit, I think we need to understand what's happening at the visits then, because for sure this is not the same as an old school physical. |
| **Theme 5:**<br>Discrimination<br>and lack of trust | [Group 2] Are we getting wellness visits as opposed to other groups getting physicals?...But what I'm saying to you is that, are we the ethnic group in Wellness visits in place of the physical?<br>[Group 3] I prefer it to be spoken as it is and be exactly spelled out what it is. They don't have a term that fits me. It be this thing, one day and that thing, the next day. Tell the truth about what it's for. Who's benefitting from this? I know I should be in the main benefit here, but I know that's not the way it is because there's a financial aspect.<br>[Group 3] Just draw the line North Side Chicago versus South Side, Chicago. Just draw the line real quick. What kind of services are they getting versus what we're not getting, or vice versa?<br>[Group 2] For example, if Northwestern gets a group of nurse practitioners together and make them part of a Wellness care group, and they want to go out, and they see the addresses that they're going to, |
|  | They may deny... So we start getting second tier treatment in our neighborhood. and it frightens me because we have a history of folks not providing great medical services to us and in our neighborhoods.<br>[Group 2] And beyond that there's a historical trepidation in the African American community related to doctors, and it can go all the way back...we retain that history to say, I don't trust doctors. |

#### Participants Value Their Health and Preventive Care

One key theme we observed in the qualitative data was that participants value their health and the potential benefits of preventive visits. Many discussed how they see preventive visits as an important aspect of protecting or maintaining their health, and that these visits may be especially beneficial for older adults. Preventive visits were also cited as a venue for communicating with primary care physicians (PCPs) and providing updates on their life and health.

#### Participants Value their Relationship with PCP

Another theme that emerged was the value that participants placed on their relationship with their PCP and continuity of care. Many participants stated that preventive visits offered an opportunity to see a trusted PCP who knew them well, and many positively mentioned the longevity of their relationship with their PCP. Relatedly, when asked about the potential to complete an AWV with a clinician who was not their PCP, participants had mixed opinions, with some citing concerns about other clinicians’ ability to address or identify clinical issues that are not standard components of the AWV.

#### Barriers to Care

Participants face barriers when scheduling and attending in-person clinical visits. Despite participants’ perceptions of AWVs’ potential benefits, they frequently reported challenges during the scheduling process. Participants described long periods of being on hold when attempting to schedule appointments via phone, needing to schedule appointments far in advance due to limited appointment availability, or being encouraged to schedule visits with another clinician besides their PCP. These barriers led participants to feel de-valued and concerned about potential health consequences associated with care delays. Participants also reported transportation-related barriers and costs to visit attendance, including parking and issues with public transportation in urban and suburban areas. Some participants stated that they scheduled multiple medical appointments on the same day to minimize the number of trips, and costs, associated with transportation to medical appointments.

#### Confusion with AWV Terminology

Although participants were familiar with preventive care in general, they had less knowledge about the specific AWV encounter and terminology. Participants had questions related to whether or not the AWV was a “physical,” which was a less ambiguous term. Some participants were not sure if they were up to date with their AWV because although they had completed a recent preventive visit, they did not know if it was technically an AWV. Many patients referenced “annual” visits in earlier adulthood and had questions about if those visits were AWVs.

One point of confusion that emerged, especially in the FQHC focus groups, was the impact of in-home visits offered by Medicare Advantage plans, which are often described with terms such as Healthy Home Visits^20^ or HouseCalls, sometimes also occurring annually with an insurance incentive.^21^ Participants enrolled in Medicare Advantage programs reported frequent contacts from insurers to promote these in-home visits, and were unsure if these encounters “counted” as their AWV, or if data collected at these visits were shared with their PCP’s office.

#### Historical Discrimination

Fifth, a recurring theme that independently emerged during multiple focus groups was historical discrimination based on race and ethnicity. Participants voiced concerns about whether there were differences in uptake of AWVs because Black and Hispanic patients may distrust whether they are receiving equitable, high-quality care. In one focus group, participants discussed the potential for suspicion about AWVs being a low-quality service only offered to disfavored populations.

## DISCUSSION

In this qualitative study of older racial and ethnic minority patients’ perceptions of Medicare AWVs, participants reported varying forms of preferred communication, including the patient portal, phone calls, and mailed letters. Key themes that emerged included the value that participants place on their health, potential benefits of preventive visits, and PCP relationships. Participants reported several barriers to AWV scheduling and completion, ranging from call center hold times to transportation costs. Additionally, participants had confusion about the meaning of an “Annual Wellness Visit,” and whether AWVs differed from other checkup visits they had previously completed. The topic of historical racism and discrimination arose during multiple focus groups.

As observed previously, focus group participants value preventive care and the potential benefits of preventive visits, but were confused by the term “Annual Wellness Visit.”^12^ This confusion may be compounded by other factors that emerged during our focus groups, such as outreach by Medicare Advantage plans to encourage patients to complete in-home exams. Additionally, though prior studies have found that AWVs with non-PCP clinicians are feasible to implement and beneficial to patients,^22–24^ this type of AWV was not necessarily desirable to current study participants, who placed a high value on time spent with a trusted PCP. Additionally, even in our focus groups at an academic health system where patients reported a high level of patient portal use, many patients reported using other modes of communication (e.g. phone calls) and expressed heterogeneous preferences for how they wanted to be contacted.

Our findings offer important insights for the design of interventions to increase AWV uptake, several of which we have incorporated in an ongoing pilot trial. As patients hold variable preferences for communication with their primary care teams, interventions are more likely to be effective if they leverage multiple forms of outreach, or use customized outreach workflows that target patients via their preferred forms of communication. Interventions should provide educational information about “Annual Wellness Visits,” the components of AWVs, potential benefits of AWVs, and highlight the fact that AWVs are recommended for *all* Medicare enrollees (not only for people from historically excluded groups). Additionally, in settings where AWVs are delivered by PCPs, this offering should be highlighted as a way to deepen patient-PCP relationships, and increase the perceived benefits of AWVs among patients who highly value encounters with their trusted PCP.

Interventions should also seek to reduce friction and costs to patients during AWV scheduling and attendance. Possible interventions to explore include promotion of virtual or telehealth AWV encounters when appropriate and acceptable to the patient, phone-based outreach and scheduling assistance by clinical staff (without wait times or putting the patient on hold), or connecting patients with services to address transportation barriers (along with parallel efforts to screen for transportation barriers). As some patients reported in our focus groups, scheduling an AWV on the same day as a second appointment with a different clinician—at the same medical campus—could also minimize transportation barriers.

This study has some notable limitations. As this study was conducted among urban, racial and ethnic minority older adult patients, our findings may not reflect the opinions and experiences of other sociodemographic groups, such as non-Hispanic Whites or non-elderly Medicare enrollees. These focus group discussions did not address the topic of virtual or telehealth AWV encounters; this approach to AWV delivery gained popularity during the early months of the COVID-19 pandemic, but then declined to only 2% of AWVs during the year 2022.^25^ Study strengths include the collection of data from patients in two organizations, in two languages, and our emphasis on patient populations known to have low rates of AWV completion, both locally and nationally.^7–10^

In conclusion, in focus groups with older racial and ethnic minority patients, participants reported heterogeneous communication preferences and that they place high value on preventive care and PCP relationships, but were confused about AWVs and how these visits differ from in-home exams offered by Medicare Advantage plans. Interventions to increase Medicare AWV uptake in at-risk populations must overcome and address these factors, patients’ barriers to visit scheduling and attendance, and any concerns related to discrimination of historically excluded groups.

## Data Availability

The authors do not currently have permission to share all data collected in this study. However, all reasonable requests will be considered and reviewed by the authors.

## Acknowledgments

Funding support was provided by the National Institute on Aging (R21AG081895). The funder had no role in the design and conduct of the study; management, analysis, and interpretation of the data; preparation, review, or approval of the manuscript; or decision to submit the manuscript for publication. Dr. Linder additionally reports grant support from the National Institute on Aging (P30AG059988, R01AG074245, P30AG024968, R01AG070054, R24AG064025, U24AG059624, U19AG065188), the National Heart, Lung, and Blood Institute (R01HL167023), the National Institute of Neurological Disorders and Stroke (U01NS105562), and the Agency for Healthcare Research and Quality (R01HS026506, R01HS029328). We wish to thank patients who served as focus group participants, as well as staff at participating practices where focus groups were held. We additionally wish to thank our partners at AllianceChicago for supporting data collection efforts during study recruitment.

